# AlphaGenome identifies a deep intronic variant in a family with PLA2G6-associated neurodegeneration: Closing the diagnostic gap in rare genetic diseases

**DOI:** 10.64898/2026.06.10.26355004

**Authors:** Sarah J Eger, Greizy López, Luisa Fernanda Gómez Navarro, Andrés Peña-Tauber, J Nicholas Cochran, Susan M Hiatt, Nancy Gelvez, Milena García-García, Samuel Lobo, Michael D Greicius, Diana Lucía Matallana, Juliana Acosta-Uribe, Kenneth S Kosik

## Abstract

A molecular diagnosis remains out of reach for a substantial subset of patients with clinically recognizable Mendelian disorders, even after comprehensive next-generation sequencing. Causal variants in non-coding regions are difficult to detect and interpret using standard pipelines. Deep intronic variants that disrupt splicing are a known but underexplored source of pathogenic alleles, and systematic tools to evaluate them at scale have only recently emerged. We aimed to resolve an incomplete genetic diagnosis in two siblings with early-onset parkinsonism, prominent neuropsychiatric features, and autonomic dysfunction consistent with *PLA2G6*-associated neurodegeneration (PLAN), an autosomal recessive condition. Prior clinical exome sequencing, genome sequencing, Multiplex Ligation-dependent Probe Amplification (MLPA), and long-read sequencing had identified only a single heterozygous *PLA2G6* missense variant, c.2132C>G (p.Pro711Arg). We used AlphaGenome to score 91 non-coding variants shared among the affected siblings and their father within 1 megabase of the *PLA2G6* locus. The deep-learning model identified an intronic variant (c.2034+355G>A) that was predicted to create a cryptic splice acceptor site that could result in inclusion of a 160-bp cryptic exon. Tissue-specific predictions indicated the aberrant splicing would be detectable in blood, confirmed by junction-spanning RNA-seq reads from an unrelated carrier. This analysis completed a compound heterozygous PLAN diagnosis nearly two decades after symptom onset and demonstrates the utility of sequence-to-function models. Systematic integration of tools like AlphaGenome into rare disease workflows offers a practical, low-barrier route to closing the diagnostic gap for patients with compelling Mendelian phenotypes and incomplete genetic diagnoses.

## Introduction

Although next generation sequencing has substantially improved rare disease diagnosis, some cases with clinically recognizable Mendelian disorders remain genetically unsolved^1^. In some cases, pathogenic alleles may reside in deep intronic regions where variants can disrupt splicing^2^, but are difficult to detect or interpret using conventional analysis pipelines. Deep learning models trained on genomic sequences, such as AlphaGenome, now make it possible to systematically evaluate non-coding variants for effects on gene expression, splicing, and other modalities, offering a practical route to provide accurate genetic diagnosis for previously unsolved cases.^3^

*PLA2G6*-associated neurodegeneration (PLAN) is a group of autosomal recessive disorders caused by pathogenic variants in *PLA2G6*, which encodes calcium-independent phospholipase A2β (iPLA2β). This enzyme has important roles in phospholipid remodeling, membrane homeostasis, mitochondrial integrity, and neuronal maintenance^4^. Historically, PLAN has been classified into three major clinical presentations: infantile neuroaxonal dystrophy (INAD), atypical neuroaxonal dystrophy (ANAD), and PARK14-associated early-onset Parkinson disease. However, evidence increasingly supports the view that PLAN represents a phenotypic spectrum with considerable variability in age of onset and clinical manifestations^5–7^.

To date, the majority of reported pathogenic *PLA2G6* variants are exonic or located at canonical intron–exon boundaries. Even so, unresolved cases continue to emerge in which only a single pathogenic coding variant is identified despite a phenotype highly suggestive of PLAN^8^, raising the possibility of a second, non-coding variant in trans^9,10^.

Here, we describe two siblings with early-onset parkinsonism consistent with PLAN in whom initial variant annotation identified only a single heterozygous missense variant in *PLA2G6*. Application of AlphaGenome revealed a deep intronic variant that was predicted to create a cryptic splice acceptor site, completing the diagnosis of compound heterozygous PLAN and demonstrating the value of sequence-to-function modeling in unsolved rare genetic diseases.

## Results

### Clinical Description

The proband and their affected sibling developed a remarkably similar clinical syndrome, with onset at between 30-35 years (Fig. 1a). Initial symptoms included gait impairment and progressive parkinsonism with bradykinesia, rigidity, and tremor, followed by dyskinesias, dystonia, postural instability, and falls. Non-motor symptoms included urinary dysfunction, severe constipation, sleep disturbance, and progressive speech impairment, which prompted an initial diagnosis of early-onset Parkinson’s disease (PD).

**Figure 1.**
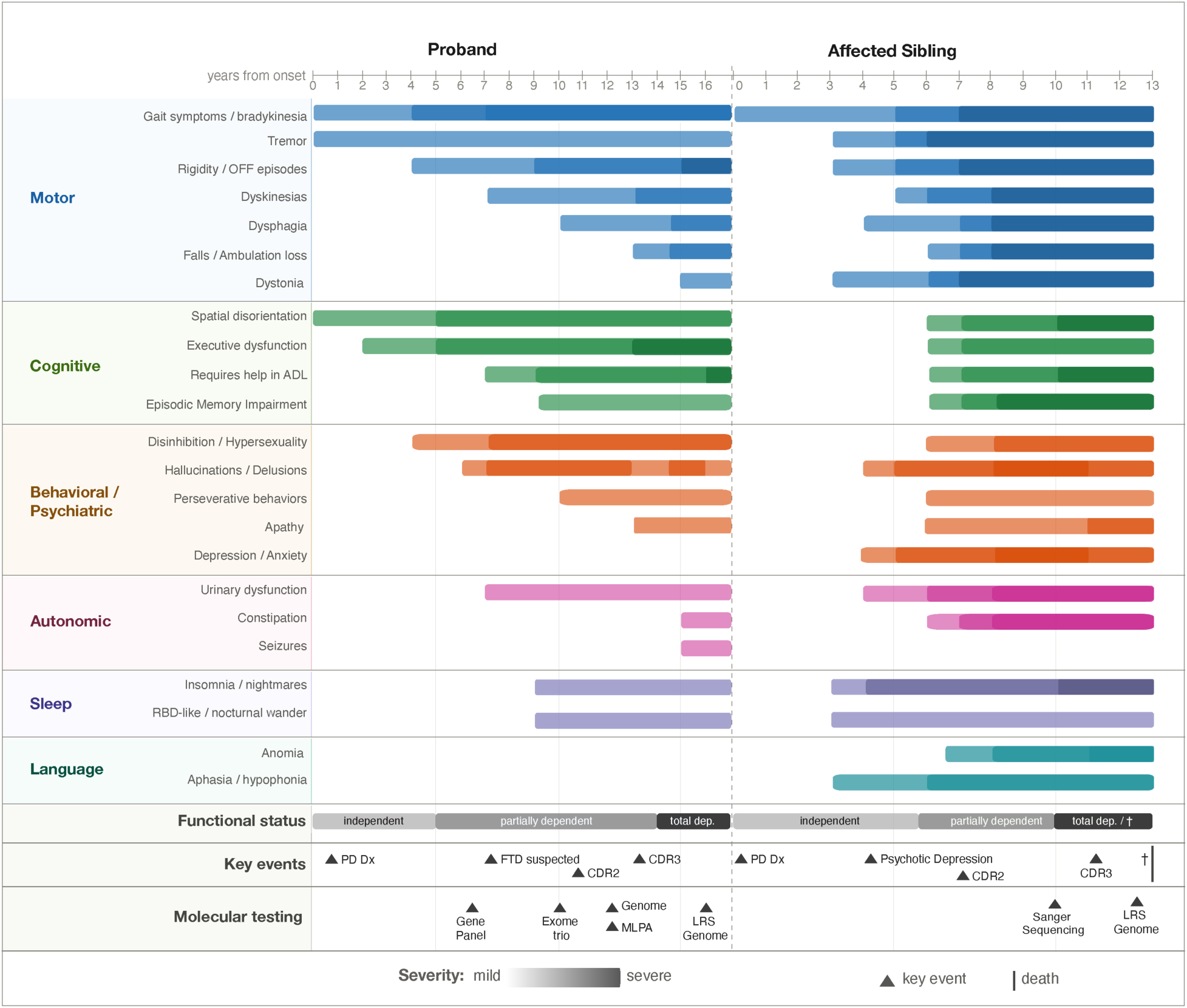
Longitudinal clinical progression of a sibling pair with *PLA2G6*-associated neurodegeneration (PLAN). Symptom timelines are aligned to years from disease onset for the proband (left) and affected sibling (right). Horizontal bars indicate the approximate duration and relative severity (light = mild; dark = severe) of symptoms across six domains: motor, cognitive, behavioral/psychiatric, autonomic, sleep, and language. Functional status is shown below as a continuous trajectory from independence to total dependence. The proband has a 17-year documented disease course with current total dependence; the affected sibling had a 13-year course ending in death. Triangular markers (▲) denote key clinical milestones, including Parkinson’s disease diagnosis, psychiatric hospitalization, and Clinical Dementia Rating stage assessments. Molecular testing modalities performed for each individual are indicated along the bottom. Symptom domain classification follows Magrinelli et al. (2021). ADL, activities of daily living; CDR, Clinical Dementia Rating; Dep., dependent; Dx, diagnosis; LRS, long-read sequencing; MLPA, multiplex ligation-dependent probe amplification; PD, Parkinson’s disease; RBD, REM sleep behavior disorder.

Both siblings demonstrated an initial motor response to dopaminergic therapy but developed prominent neuropsychiatric symptoms within five years of motor symptom onset. The proband exhibited behavioral disinhibition with hypersexuality, hyperphagia, and grandiose delusions. The affected sibling developed a more apathetic phenotype characterized by irritability, flat affect, and perseverative behaviors. Visual hallucinations occurred in both siblings. This rapid neuropsychiatric progression was atypical for PD, prompting consideration of alternative diagnoses including frontotemporal lobar degeneration with parkinsonism or Lewy body disease.

Neuroimaging findings were nonspecific with respect to the underlying neurodegenerative etiology. The proband had generalized cerebral atrophy, with sulci widening and increased ventricular volume. Susceptibility-Weighted Imaging (SWI) showed mild hypointensity of the basal ganglia consistent with mineral deposition (Supplementary Fig. 1).

With progression of neuropsychiatric symptoms, both siblings’ cognitive decline advanced from executive dysfunction and memory impairment to loss of independence and dementia.

### A genetic diagnosis is suspected

Given the early age at onset and the phenotypic concordance between siblings, a genetic etiology was suspected. Seven years after symptom onset, the proband underwent targeted gene panel sequencing for PD. Variant annotation identified 30 coding variants; following curation, two variants of uncertain significance (VUS) in *FBXO7* (NM_012179.3:c.1546G>C, p.(Asp516His), rs34316445), and *PLA2G6* (NM_003560.4:c.2132C>G, p.(Pro711Arg), rs199784053) were detected (Supplementary Table 1). Notably, the proband was homozygous for a *LRRK2* haplotype carrying variants p.Asn551Lys and p.Lys1423Lys, which has been reported to be protective for PD^11,12^. The *PLA2G6* p.Pro711Arg variant was classified as a VUS in ClinVar (VCV001381529.6) at the time of reporting; however, it had been previously linked to the INAD form of PLAN in a compound heterozygous case.

Trio exome sequencing, performed two years later, did not identify pathogenic variants in dementia associated genes, nor additional protein altering variants in the coding and flanking regions (±8 bp) of *FBXO7* and *PLA2G6,* and confirmed these variants to be inherited from the father and mother, respectively (Supplementary Table 2). Variant curation at the time classified the *PLA2G6* p.Pro711Arg variant as likely pathogenic, while *FBXO7* p.Asp516His remained as VUS.

Although only one pathogenic variant had been identified, the clinical picture strongly supported a PLAN diagnosis. The combination of early-onset parkinsonism, early dyskinesias, prominent psychiatric and behavioral symptoms, autonomic dysfunction, and progressive cognitive decline in both affected siblings matched the established phenotype of late-onset PLAN^7^. Both parents were older than 75 years and unaffected (Fig. 2), consistent with autosomal recessive inheritance, raising the possibility that a second pathogenic allele inherited from the father and located outside the exome capture regions, remained unidentified. To investigate this, three additional studies were performed four years after trio exome sequencing. Clinical whole genome sequencing of both siblings did not identify non-coding pathogenic variants. Multiplex Ligation-dependent Probe Amplification (MLPA) of *PLA2G6*, performed to detect structural variants (SVs) previously linked to PLAN^13,14^, revealed no deletions or duplications at the locus. Spinocerebellar ataxia triplet expansion analysis was also normal.

**Figure 2.**
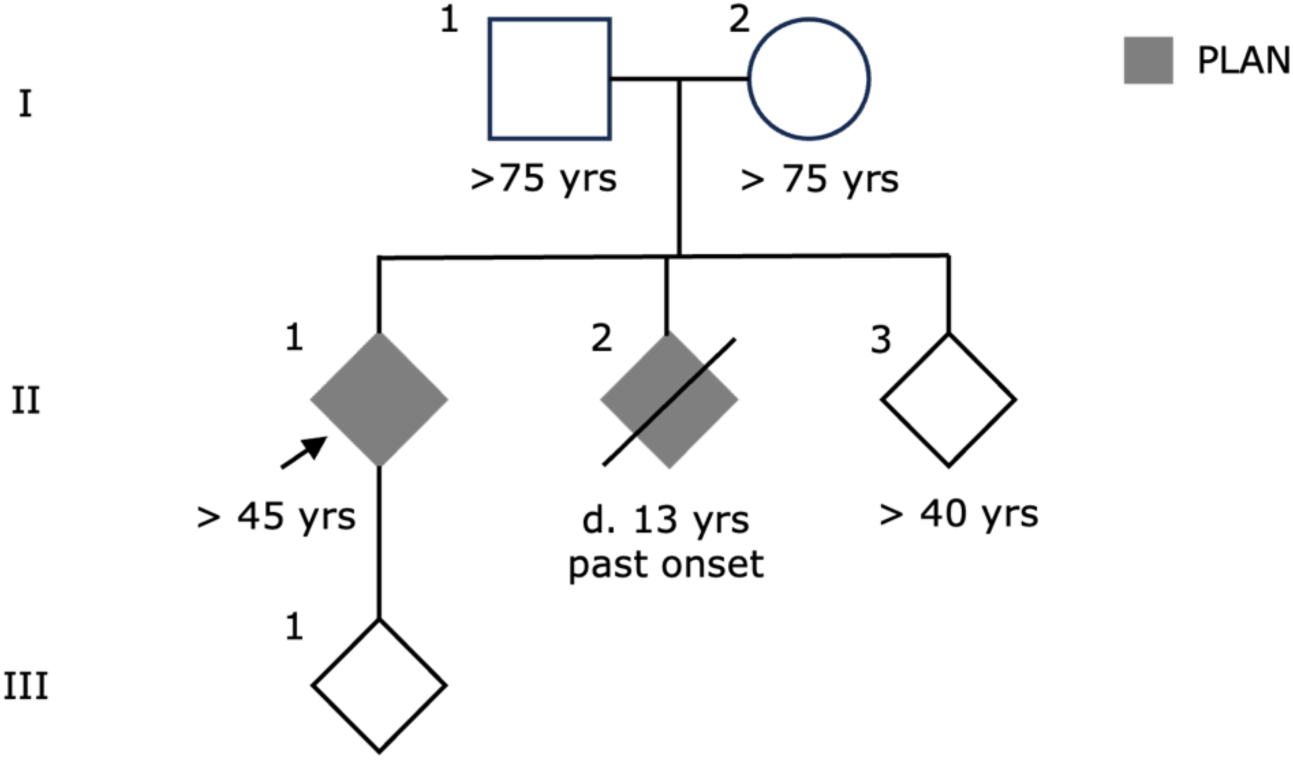
Three-generation pedigree of the family. Generation I comprises an unaffected couple (I-1, age >75; I-2, age >75). Generation II includes three offspring: the proband (II-1, arrow) and an affected sibling (II-2, deceased) both with PLA2G6-associated neurodegeneration (PLAN, shaded), and an unaffected sibling (II-3, >40 ). The proband (II-1) has one child (III-1).

#### Searching for the second variant

To assess copy number variations and tandem repeat expansions in other neurodegeneration associated genes, we performed PacBio long-read whole genome sequencing for both the proband and the affected sibling. SV calling with pbsv and annotation with annotSV and SvAnna did not detect any SVs in *PLA2G6* Inspection of the locus in IGV showed no evidence of coding variation that could explain loss of function of the paternal haplotype. Tandem repeat expansions were also called using TRGT. No expansions were found in *C9orf72*.

To identify candidate non-coding variants, we used AlphaGenome to evaluate all variants shared by the proband, affected sibling, and father within 1 Mb of *PLA2G6*. This yielded 91 variants for AlphaGenome annotation (Supplementary Table 3). We scored variants across all tissues using AlphaGenome’s recommended scorers for four genomic modalities: RNA-seq, splice site probability, splice site usage, and splice junction. Analysis ran in 3 minutes and 21 seconds on eight NVIDIA H100 GPUs.

AlphaGenome assigned scores to 91 variants in the *PLA2G6* region (Supplementary Table 4). One variant, chr22:38115172:C>T (rs147795054), corresponding to *PLA2G6* NM_003560.4:c.2034+355G>A, was a clear outlier, as it comprised the top 735 out of 264,566 AlphaGenome scores across all tissues, and was the only variant with scores above 2, with a maximum score of 8.26. On manual inspection, this was driven by splice junction scores, in which it was a step above all other variants examined (Fig. 3a). The splice junction score reflects the maximum absolute log-fold change in predicted junction counts across splice site pairs, capturing changes in both splice site usage and splicing efficiency. AlphaGenome predicted that the variant creates a cryptic splice acceptor AG site that pairs with a pre-existing intronic GT donor, defining a 160-bp cryptic exon within intron 14 (Fig. 3b). Inclusion of this cryptic exon would cause a frameshift starting at residue position 679 and ending in a new predicted stop codon a further 146 residues downstream (protein sequence included as supplementary material 1). *PLA2G6*, NM_003560.4:c.2034+355G>A also scored highly with SpliceAI with an acceptor gain delta score of 0.95 and the same predicted splicing consequence, but was tissue-agnostic^15,16^.

**Figure 3.**
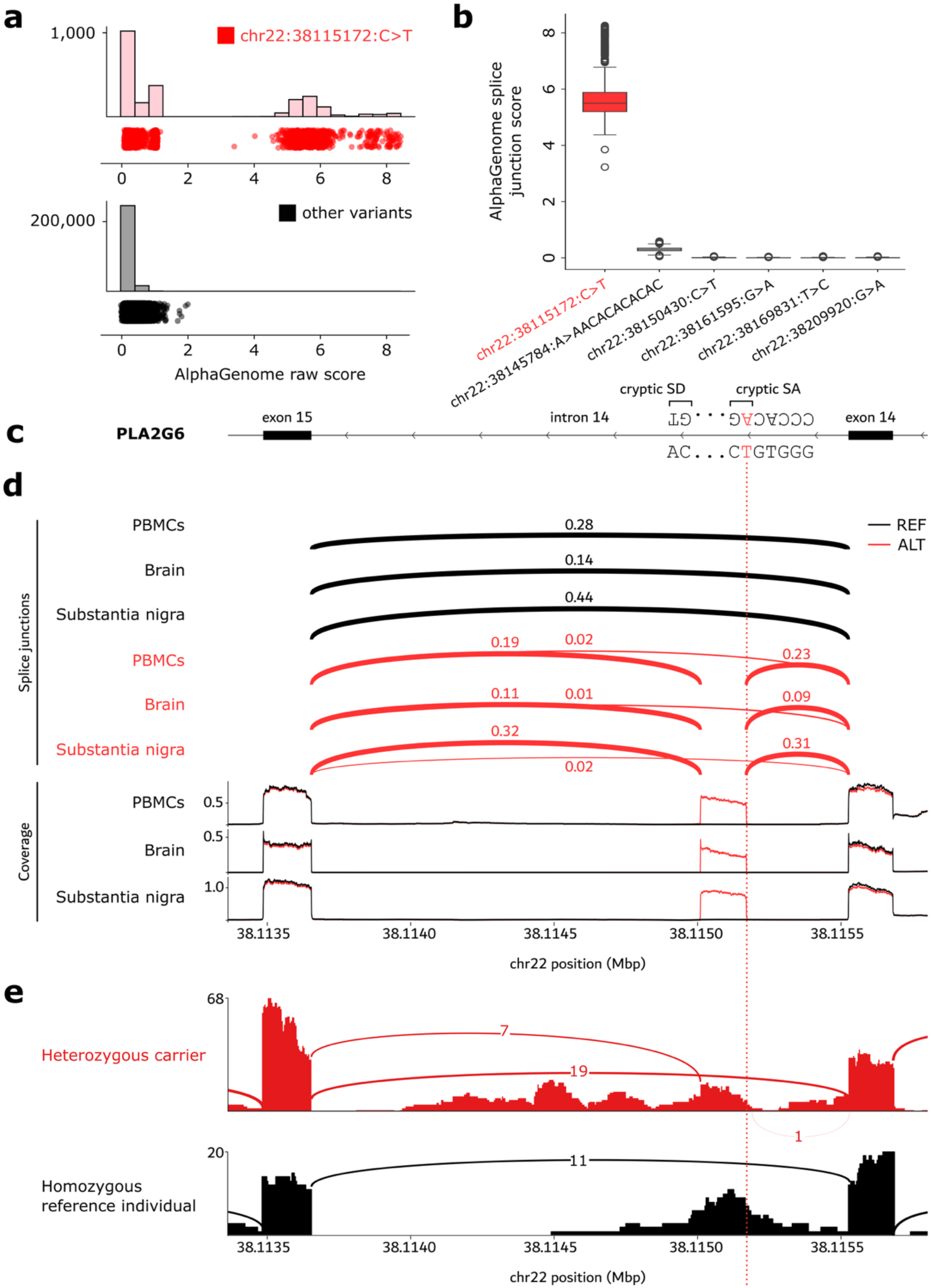
A deep intronic *PLA2G6* variant causes cryptic exon inclusion, accurately predicted by AlphaGenome. **(a)** AlphaGenome scores from recommended scorers in all tissues, for variants in a 1Mbp region centered on *PLA2G6*. **(b)** AlphaGenome raw splice junction scores for six variants overlapping the *PLA2G6* genomic locus. Scores represent the maximum absolute log-fold change in predicted junction counts across all splice site pairs. **(c)** Schematic of *PLA2G6* intron 14 illustrating the variant-created cryptic splice acceptor (SA) site and a pre-existing cryptic splice donor (SD) site that together define a 160-bp cryptic exon. Sequence context at each site is shown. **(d)** AlphaGenome-predicted splice junctions (top) and RNA-seq coverage tracks (bottom) for the reference (REF, black) and alternate (ALT, red) alleles across PBMCs, brain, and substantia nigra. Numbers indicate predicted junction usage proportions. The dashed red line marks the position of the cryptic SA site. **(e)** Sashimi plots of RNA-seq data from a heterozygous carrier (red) and a homozygous reference individual (black), showing read coverage and split-read counts supporting canonical and cryptic splice junctions. Numbers on arcs indicate supporting read counts. SA, splice acceptor; SD, splice donor; PBMC, peripheral blood mononuclear cell.

Visualization of predicted variant effects showed that in both brain and substantia nigra, tissues most relevant to PLAN, the canonical exon 15-to-exon 14 junction would be partially reduced. Additionally, two new junctions flanking the cryptic exon could emerge and transcripts could include the cryptic exon (Fig. 3c). Notably, equivalent effects were predicted in peripheral blood mononuclear cells (PBMCs), suggesting the splicing defect may be detectable in blood-derived RNA from carriers.

To test this, we identified an unrelated heterozygous carrier of NM_003560.4:c.2034+355G>A with available whole blood RNA-seq from AMP-PD. This individual showed junction-spanning reads connecting the cryptic exon to both flanking exons. A homozygous reference individual showed only the canonical junction with no cryptic junction reads (Fig. 3d). Together, these results support AlphaGenome’s and SpliceAI’s predictions and establish that this deep intronic variant causes cryptic exon inclusion detectable in carrier RNA.

Interestingly, in AMP-PD database, we found two individuals who were homozygous for the intronic variant. One had Lewy Body Dementia at an age older than 75, while another was a healthy control at an age older than 65. Across gnomAD, All of Us, and BRAVO/Topmed, we found a total of 24 homozygotes. Therefore, it appears unlikely that being homozygous for this variant necessarily results in disease.

#### Genetic diagnosis

Identification of causal variants on both alleles enabled a complete genetic diagnosis for the family. Segregation analysis showed that the affected siblings inherited the missense variant, NM_003560.4 c.2132C>G (p.Pro711Arg), from their mother and the deep intronic variant, NM_003560.4:c.2034+355G>A, from their father. The unaffected sibling carried only the deep intronic variant. The proband’s offspring underwent genetic counseling and testing to determine carrier status. Sanger sequencing confirmed they carried only the missense variant (Supplementary Figure 2), indicating they are unlikely to develop PLAN.

## Discussion

Many patients with rare Mendelian disorders either never receive a genetic diagnosis or receive one too late for therapeutic intervention or timely genetic counseling of at-risk relatives. This diagnostic gap stems partly from the heavy bias of curated disease databases toward coding single nucleotide variants. Long-read sequencing has expanded the diagnostic toolkit by resolving structural variants and tandem repeat expansions, yet a subset of cases remain unsolved. In many of these cases, the causal variant likely resides in intronic sequences that standard tools are poorly equipped to evaluate. Deep learning models trained on genomic sequences can predict the splicing consequences of non-coding variants, providing a practical path to diagnosis in cases where standard approaches fall short.

In the present study, standard clinical exome sequencing and genome-wide analysis identified only a single heterozygous missense *PLA2G6* variant. MLPA and long-read sequencing detected no structural variation in the gene, leaving the family without a complete molecular diagnosis for nearly two decades after the proband’s symptom onset, and three years after the affected sibling died. Applying AlphaGenome to score non-coding variants shared by both siblings, identified a deep intronic *PLA2G6* variant predicted to create a cryptic splice acceptor site and introduce a frameshift through inclusion of a 160-bp cryptic exon, completing the compound heterozygous diagnosis. The missense variant is most likely a gain-of-function allele, whereas the deep intronic variant may act through loss of function; this effect would not be pathological in isolation but, in the absence of a normal second allele, produces disease. This represents an alternative route to compound heterozygosity, in which one allele carries a coding gain-of-function variant and the other a non-coding loss-of-function variant.

Several observations argue that the deep intronic variant is not independently pathogenic but requires a more damaging variant in trans to cause disease. First, population-scale analyses have not found *PLA2G6* variants to be enriched in Parkinson’s disease cases compared with controls^17^. Second, the variant occurs at notably higher frequency in Finnish populations (3.5% in gnomAD v4.1.1), and we found at least one healthy homozygote. Taken together, these findings suggest the variant may require a second, more pathogenic allele in compound heterozygosity to cause disease or exhibit reduced penetrance.

Whole-genome sequencing returns many non-coding variants with no clear way to prioritize them. Even when a candidate is identified, standard *in silico* tools offer limited guidance on where to look for functional confirmation. RNA sequencing can resolve this, but it is costly and requires knowing which tissue to sample. Two PLAN cases illustrate the problem. Cavestro et al. resolved a similar half-solved PLAN diagnosis by sequencing patient RNA directly, which revealed an aberrant transcript and guided them back to the causal variant^9^. This was an effective approach, but one that requires investing in RNA analysis before any candidate variant is in hand. Borja et al. identified a different deep intronic variant in *PLA2G6*^10^. SpliceAI predicted a cryptic splice acceptor gain, but its tissue-agnostic score left the authors unable to determine whether the effect would be detectable in blood and thus which tissue to sequence. AlphaGenome addresses both problems: it narrows candidate variants before RNA analysis is undertaken, and its tissue-specific predictions allow tissue selection to precede RNA sequencing. Both prior studies identified deep intronic variants in intron 14 predicted to alter splicing, suggesting this region of *PLA2G6* may be enriched for pathogenic variants and warrants close examination in genetically unresolved PLAN cases.

The main limitation of our study is the absence of RNA sequencing from the affected individuals and their variant-carrier father. Future work should quantify mRNA levels of the introduced cryptic exon to directly confirm aberrant splicing and determine whether nonsense-mediated decay reduces transcript abundance. As a result, cryptic exon inclusion could not be confirmed directly in the disease-relevant carriers. Validation instead relied on an unrelated carrier whose blood RNA-seq showed junction-spanning reads consistent with AlphaGenome’s prediction, but this does not substitute for functional confirmation within the family. Neither does it allow quantification of aberrant transcript levels or assessment of nonsense-mediated decay.

As a single-family report, the findings cannot be generalized without replication in independent cases. The diagnostic approach applied here, scoring non-coding candidates with AlphaGenome across affected family members, could be extended to other unsolved PLAN cases carrying only one identified coding allele. More broadly, this approach applies to any Mendelian phenotype where a second pathogenic allele remains unidentified. Now that both alleles have been identified, genetic counseling for at-risk relatives is an immediate clinical priority.

Many patients with clinically recognizable Mendelian disorders go undiagnosed because causal variants fall outside the reach of standard analysis pipelines; this case demonstrates that deep learning models trained on genomic sequence can find them. Clinical exome sequencing, whole-genome sequencing, MLPA, and long-read structural variant analysis each failed to complete the molecular diagnosis in this family. Applying AlphaGenome to score non-coding candidates resolved it in minutes, nearly two decades after symptom onset. The practical barrier to adopting this approach is low: it requires only a list of candidate variants shared across affected individuals and access to a GPU. For patients with a compelling Mendelian phenotype and an incomplete genetic diagnosis, integrating sequence-to-function models into clinical variant curation is the most direct route to closing this gap.

## Methods

### Clinical assessment

The two affected siblings underwent standardized clinical, neurological, and neuropsychological examination following the protocol of the ReDLat study^18^. Retrospective neurological examinations records were used to characterize the progression of symptoms. Brain MRI was performed on a 1.5-Tesla system using spin-echo radiofrequency pulse sequences with T1- and T2-weighted protocols, including axial fluid-attenuated inversion recovery (FLAIR) and axial susceptibility-weighted imaging (SWI).

All participants or their legally authorized proxies signed an informed consent approved by the institutional review board (IRB) of the Pontificia Universidad Javeriana - Hospital Universitario San Ignacio (FWA00001113).

### Clinical genetic sequencing

The proband underwent targeted gene panel sequencing encompassing 19 genes associated with Parkinson’s disease. Exonic regions were captured using the TruSight One Sequencing Panel and sequenced on the NextSeq 500 platform (Illumina, 2×150bp paired-end), achieving >20X depth in 89.62% of the coding region. Variants were annotated and cross-referenced against publicly available variant databases and in silico pathogenicity prediction tools.

Whole-exome sequencing was performed on the proband and both parents (trio analysis). Coding regions and flanking sequences were enriched using Agilent solution-based capture and sequenced on the Illumina HiSeq/NovaSeq system. Coverage of target regions at a minimum of 30 high-quality reads per base was 93.91%, 95.05%, and 90.54% in the proband, mother, and father, respectively. Variants were annotated and filtered for SNVs and small indels in coding regions and flanking sequences (±8 bp) with a MAF <1.5% (GnomAD). In silico pathogenicity was assessed using Mutation Taster, FATHMM, Mutation Assessor, SIFT, FATHMM-MKL, LRT, and PROVEAN, with consensus thresholds defined as: 100% = pathogenic/benign; ≥75% = probably pathogenic/benign; <75% or no prediction = inconsistent. Priority was assigned to 32 genes associated with Parkinson’s disease and 18 genes associated with dementia.

Multiplex Ligation-dependent Probe Amplification (MLPA) analysis was performed using Probemix P120-B2 (MRC Holland) to capture *PLA2G6* (NM_003560.2; chr.22): all exons (except exons 7 and 17), intron 6, and 3’UTR.

Spinocerebellar ataxias triplet expansion analysis was done to identify amplification of the clinically significant repetitive regions of *ATN1, ATXN1, ATXN2, ATXN3, CACNA1A, ATXN7*.

Whole genome sequencing (WGS) of the affected sibling was performed by GencellPharma, Colombia. Variants were classified and reported using the Golden Helix VarSeq analysis workflow. WGS of the unaffected sibling, mother, and father was performed as described in Acosta-Uribe et al. (2026)^18^. Kinship coefficients for all family members matched the expected values based on the pedigree.

For the proband’s child, Sanger sequencing was performed for both *PLA2G6* variants by GENEWIZ from Azenta and analyzed using the Ugene software v53.1.

See Supplementary methods for a detailed description of the extensive genetic sequencing and curation processes.

### Long-read sequencing

Long-read genome sequencing was performed by HudsonAlpha, as previously described^19^. Briefly, genomic DNA from the proband was sequenced using PacBio HiFi (CCS) chemistry on a Sequel IIe instrument (Pacific Biosciences) with a targeted coverage of 30x. HiFi reads were aligned to the GRCh38 no-alt reference using pbmm2 (https://github.com/PacificBiosciences/pbmm2). SNVs and indels were called with DeepVariant^20^ and phased using WhatsHap^21^. Structural variants were identified with pbsv (https://github.com/PacificBiosciences/pbsv). Variant calls were annotated using an in-house analysis pipeline incorporating gnomAD population frequencies and standard clinical variant-interpretation resources. Structural variants were additionally filtered using bcftools and prioritized using SvAnna^22^. Repeat expansions were assessed in a curated set of disease-associated tandem-repeat loci using pbsv calls, IGV visualization, and TRGT/TRVZ^23^ (GRCh38 reference, BED-based repeat catalog).

### AlphaGenome annotation

To assess non-coding variation, we used the AlphaGenome research repository, version 0.2.0, available at https://github.com/google-deepmind/alphagenome_research^3^. Each variant was represented as a ‘genome.Variant object’ and centered within a 1 Mb genomic interval using ‘reference_interval.resizè. Variant effects were predicted by calling ‘model.score_variant’ with the full set of recommended scorers (‘RECOMMENDED_VARIANT_SCORERS’), spanning four genomic modalities: RNA-seq, splice sites, splice site usage, and splice junctions. Predictions were run for all available human tissues. Variants were sorted by raw score across tissues and modalities, and outliers were selected for manual review. The top-scoring candidate was selected for downstream validation.

## Data and code availability

All anonymized data not published here will be made available upon request from any qualified investigator. Code is available as supplementary material 2.

## Author contributions

Conceptualization, S.J.E., J.N.C., M.D.G., J.A-U., K.S.K.

Clinical assessment, G.L.L., M.G., D.L.M., J.A-U.,

Genetic sequencing, G.L.L., L.F.G.N., J.N.C., S.M.H., N.G.,

investigation, S.J.E., G.L.L., L.F.G.N., A.P-T., S.L.,

supervision, J.N.C., M.D.G., D.L.M., J.A-U., K.S.K.

writing – original draft, S.J.E., J.A-U. writing – review & editing, all authors.

## Data Availability

All anonymized data not published here will be made available upon request from any qualified investigator.

## Acknowledgements

We thank the family members who participated in this study. We acknowledge Russell O. Kosik for reviewing the brain MRI for the patients.

We acknowledge the support of the Alzheimer’s Association (SG-20–725707); Rainwater Charitable Foundation–Tau Consortium; Bluefield Project to Cure Frontotemporal Dementia; and the Global Brain Health Institute. This work was supported by American Heart Association Grant #26PRE1545631 / Sarah Eger / 2026. We acknowledge the generous support of an anonymous donor.

Data used in the preparation of this article were obtained from the Accelerating Medicine Partnership® (AMP®) Parkinson’s Disease (AMP PD) and Parkinson’s Disease & Related Disorders (AMP PDRD) Knowledge Platform. For up-to-date information on the study, visit https://www.amp-pdrd.org. The AMP® PD program is a public-private partnership managed by the Foundation for the National Institutes of Health and funded by the National Institute of Neurological Disorders and Stroke (NINDS) in partnership with the Food and Drug Administration (FDA), National Institute on Aging (NIA), Aligning Science Across Parkinson’s (ASAP) initiative; Celgene Corporation, a subsidiary of Bristol-Myers Squibb Company; GlaxoSmithKline plc (GSK); The Michael J. Fox Foundation for Parkinson’s Research (MJFF); AbbVie Inc.; Pfizer Inc.; Sanofi US Services Inc.; and Verily Life Sciences LLC.

## Declaration of generative AI and AI-assisted technologies in the writing process

During the preparation of this work the authors used Claude and ChatGPT to improve grammar, syntax and readability. After using these tools, the authors reviewed and edited the content as needed and take full responsibility for the content of the publication

