## Supplementary Figures, materials, methods for "AlphaGenome identifies a deep intronic variant in a family with PLA2G6-associated neurodegeneration: Closing the diagnostic gap in rare genetic diseases"

**Supplementary Table 1.** Candidate variants assessed in the exome sequencing of the proband.

**Supplementary Table 2.** Candidate variants assessed in the exome trio sequencing.

**Supplementary Table 3.** 91 variants within 1 Mbp of PLA2G6.

**Supplementary Table 4.** AlphaGenome variant scores.

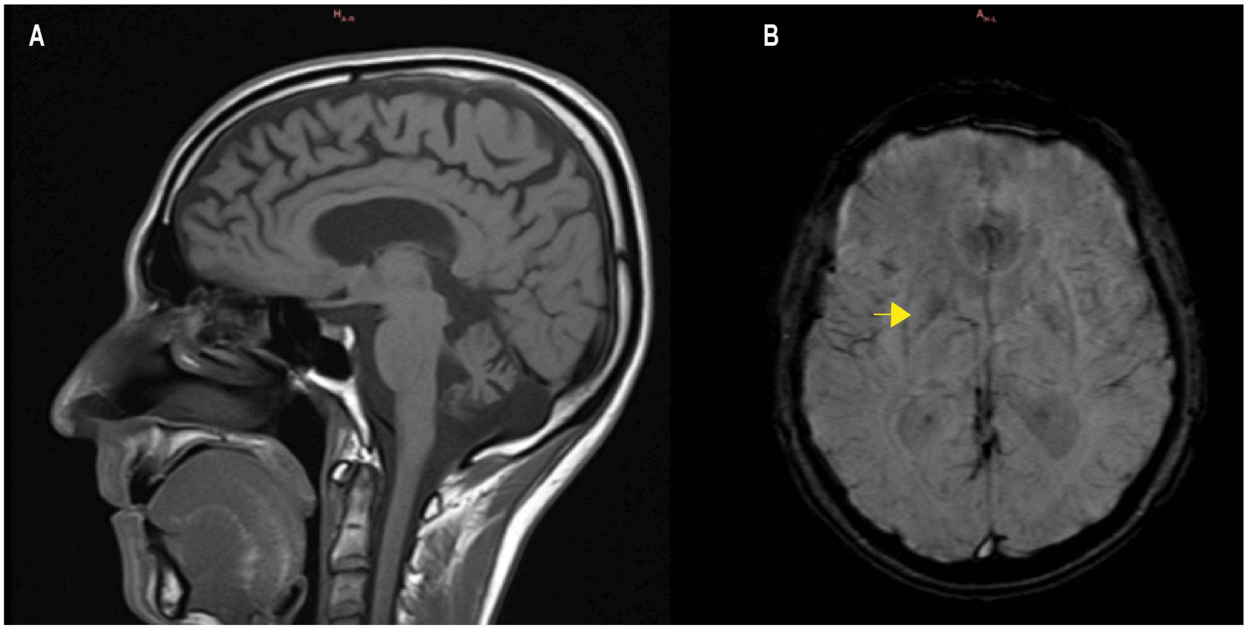


**Supplementary Figure 1.** Brain MRI **(A)** T1-weighted sagittal image shows widened cortical sulci, indicative of cortical atrophy. **(B)** Axial Susceptibility-weighted imaging (SWI) demonstrates hypointense signal in the basal ganglia, consistent with mineral deposition.

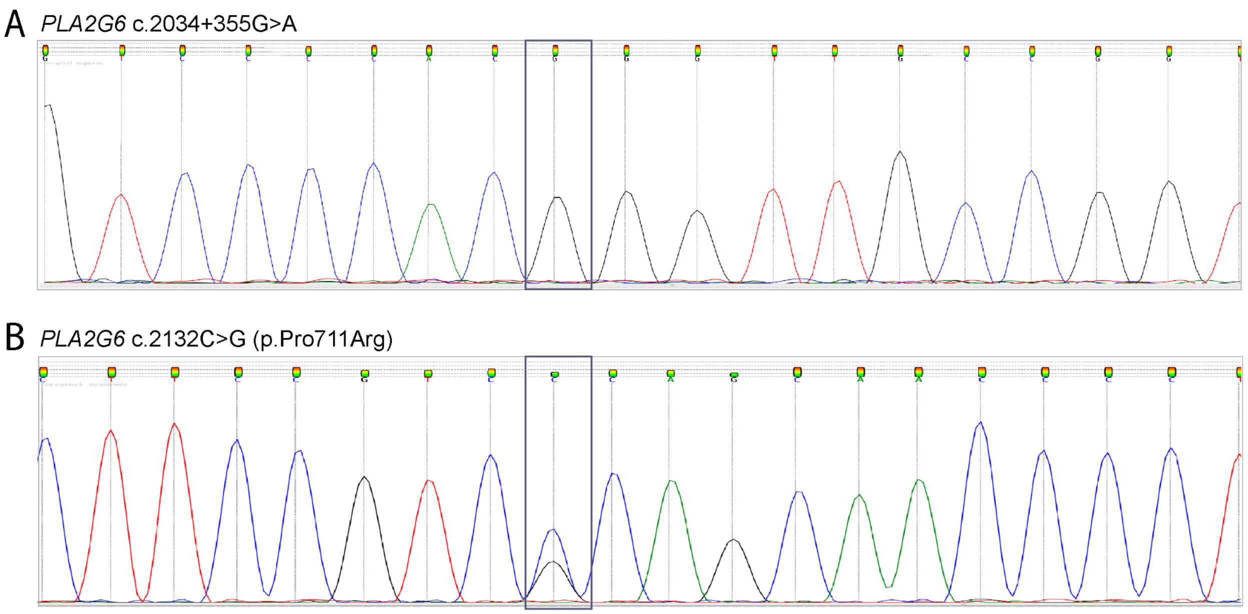


**Supplementary Figure 2.** Sanger sequencing of the proband's child. **(a)** Chromatograms for the intronic variant NM_003560.4:c.2034+355G>A, showing a G:G genotype and **(b)** for the missense variant NM_003560.4 c.2132C>G (p.Pro711Arg), showing a C:G genotype.

MQFFGRLVNTFSGVTNLFSNPFRVKEVAVADYTSSDRVREEGQLILFQNT

PNRTWDCVLVNPRNSQSGFRLFQLELEADALVNFHQYSSQLLPFYESSPQ

VLHTEVLQHLTDLIRNHPSWSVAHLAVELGIRECFHHSRIISCANCAENE

EGCTPLHLACRKGDGEILVELVQYCHTQMDVTDYKGETVFHYAVQGDNSQ

VLQLLGRNAVAGLNQVNNQGLTPLHLACQLGKQEMVRVLLLCNARCNIMG

PNGYPIHSAMKFSQKGCAEMIISMDSSQIHSKDPRYGASPLHWAKNAEMA

RMLLKRGCNVNSTSSAGNTALHVAVMRNRFDCAIVLLTHGANADARGEHG

NTPLHLAMSKDNVEMIKALIVFGAEVDTPNDFGETPTFLASKIGRLVTRK

AILTLLRTVGAEYCFPPIHGVPAEQGSAAPHHPFSLERAQPPPISLNNLE

LQDLMHISRARKPAFILGSMRDEKRTHDHLLCLDGGGVKGLIIIQLLIAI

EKASGVATKDLFDWVAGTSTGGILALAILHSKSMAYMRGMYFRMKDEVFR

GSRPYESGPLEEFLKREFGEHTKMTDVRKPKVMLTGTLSDRQPAELHLFR

NYDAPETVREPRFNQNVNLRPPAQPSDQLVWRAARSSGAAPTYFRPNGRF

LDGGLLANNPTLDAMTEIHEYNQDLIRK**VAGAVVEDREASAKRPGGSERA**

**PWNRGPILRPLGTEQCRISDLEEEQQPSLCRGSGQQGEETLHRCLPGDRE**

**VPTSACDLCGCLPSQQPLGAGQDCFWGQGTGQDGGGLLHGSRRAGCGPGT**

**GLVRDGRHPVLQIEPPAGDGHHAGX**

**Supplementary Material 1.** Predicted protein sequence for transcript including the intronic variant NM_003560.4:c.2034+355G>A

**Supplementary Material 2.** Python script for AlphaGenome variant scoring.

**Supplementary Methods**

1. **Gene Panel:**

The proband had a gene panel performed of the genes associated with Parkinson's disease, including *ADH1C, ATP13A2, DNAJC6, FBXO7, GBA, GIGYF2, HTRA2, LRRK2, MAPT, PARK2, PARK7, PINK1, PLA2G6, SLC6A3, SNCA, TAF1, TBP, UCHL1, VPS35*. 89.62% of the coding region of the genes in this panel had a depth greater than 20X.

Capture of exonic regions of interest using the TruSight One Sequencing Panel kit and generation of paired-end libraries. Ultra-sequencing (2×150bp) on the NextSeq 500 sequencing system platform (Illumina).

Bioinformatic analysis of the data using HD Genome One Research Edition software by Dreamgenics (version ImegenExomePipeline_23Sep2015):

- SNV sensitivity >99%
- Indel sensitivity (up to 18 bp) >97%
- Specificity >99.5%

Variants were considered for changes with a read depth >20 and a frequency above 30%.

Detected variants were cross-referenced against various databases and *in silico* prediction programs.

Databases Consulted:

Database of Single Nucleotide Polymorphisms (dbSNP: http://www.ncbi.nlm.nih.gov/SNP/)

1000 Genomes (http://www.1000genomes.org/)

NHLBI Exome Sequencing Project (ESP: http://evs.gs.washington.edu/EVS/)

Exome Aggregation Consortium (ExAC: http://exac.broadinstitute.org/)

Clinical Variant (http://www.ncbi.nlm.nih.gov/clinvar/)

The Human Gene Mutation Database Professional (HGMD: http://www.hgmd.org/)

IMEGEN proprietary database

In Silico Prediction Programs:

SIFT (http://sift.jcvi.org/)

PolyPhen (http://genetics.bwh.harvard.edu/pph2/)

Mutation Taster (http://www.mutationtaster.org/)

PROVEAN human genome variants (http://provean.jcvi.org/genome_submit_2.php)

LTR, SVM, FATHMM, GERP++ score

Sequencing and variant curation performed by DNA Alliance in Madrid, Spain in 2016.

2. **Exome Trio:**

A whole-exome analysis was performed on the three individuals described (father, mother, and son). The following genes were given priority given the patient's phenotype.

*ATP13A2, ATP1A3, C19ORF12, CHCHD2, DCTN1, DNAJC6, FBXO7, FTL, GBA, GCH1, GRN, LRRK2, MAPT, PANK2, PARK2, PARK7, PINK1, PLA2G6, PRKRA, RAB39B, SLC30A10, SLC39A14, SLC6A3, SNCA, SPG11, SPR, SYNJ1, TAF1, TH, VPS13C, VPS35* (Parkinson's Disease)

*APOE, APP, CHCHD10, CHMP2B, CSF1R, GRN, ITM2B, MAPT, NOTCH3, PRNP, PSEN1, PSEN2, SQSTM1, TARDBP, TBK1, TREM2, UBQLN2, VCP* (Dementia)

Coding regions and their flanking regions were enriched using Agilent solution-based technology and sequenced on the Illumina HiSeq/NovaSeq system. Illumina bcl2fastq2 was used to analyze sequence reads. Adapter trimming was performed using Skewer. Trimmed sequences were mapped to the human reference genome (hg19) using Burrows-Wheeler Aligner. Variants were annotated based on various internal and external databases.

Only variants (SNVs/small Indels) in the coding region and flanking regions (±8 bp) with a minor allele frequency (MAF) <1.5% were evaluated. Minor Allelic Frequency (MAF) values were taken from public databases (e.g. GnomAD). In-silico prediction of pathogenicity was classified based on results from Mutation Taster, FATHMM, Mutation Assessor, SIFT, FATHMM-MKL coding, LRT, and PROVEAN according to the following criteria: 100% consensus = pathogenic/benign; ≥75% consensus = probably pathogenic/benign; <75% consensus or no possible prediction = inconsistent. To evaluate the consequences of splicing variants, two algorithms (Jian et al., 2014, PMID: 25416802) were applied.

Variants found in the patient and in the patient's parents were compared and filtered for four scenarios: *de novo* variant in the patient, compound heterozygous patient, homozygous patient with heterozygous parents, and hemizygous patient with heterozygous mother for X-chromosome variants.

In this case, 93.91%, 95.05%, and 90.54% of target regions were covered by a minimum of 30 high-quality reads per base in the patient, mother, and father, respectively.

Sequencing and variant curation performed by Gencell Genética Avanzada in Bogotá, Colombia in 2018.

3. **Amplification of repetitive regions:**

Amplification of the clinically significant repetitive regions of *ATN1 (DRPLA), ATXN1 (SCA1), ATXN2 (SCA2), ATXN3 (SCA3), CACNA1A (SCA6), ATXN7 (SCA7)* was performed by fluorescent PCR.

- (CAG)n in exon 8 of the *ATXN1* gene (chr.6): normal alleles: 6–44 repeats (<35 normal alleles; 36–44 normal alleles with CAT interruptions), intermediate alleles: 36–38 repeats, full mutation alleles (expansion): >39 repeats. Reference sequence: NM_000332.3.
- (CAG)n in exon 1 of the *ATXN2* gene (chr.12): normal alleles: ≤31 repeats; intermediate alleles: 32 repeats; full mutation alleles (expansion): ≥33 CAG repeats. Reference sequence: NM_002973.3.
- (CAG)n in exon 10 of the *ATXN3* gene (chr.14): normal alleles: 12–44 repeats; full mutation alleles (expansion): ≈60–87 repeats. Reference sequence: NM_004993.5.
- (CAG)n in exon 47 of the *CACNA1A* gene (chr.19): normal alleles: ≤18 repeats; intermediate alleles: 19 repeats; full mutation alleles (expansion): 20–33 repeats. Reference sequence: NM_001127221.1.
- (CAG)n in exon 4 of the *ATXN7* gene (chr.3): normal alleles: ≤27 repeats (to date, no normal allele with more than 19 repeats has been reported; therefore, no data are available on the clinical significance of alleles between 19 and 27 repeats); premutation alleles: 28–33 repeats; pathological alleles with reduced penetrance: 34–36 repeats; pathological alleles with full penetrance: >36 repeats. Reference sequence: NM_000333.3.

- [(CTA-TAG)n(CTG-CAG)n] in exon 5 of the *ATXN8OS/ATXN8* gene (chr.13): normal alleles: 15–50 repeats; alleles of questionable significance: 50–70 repeats [(CTA·TAG)n(CTG·CAG)n]; full mutation alleles (expansion): >80 repeats. Reference sequence: NR_002717.2.
- (ATTCT)n in intron 9 of the *ATXN10* gene (chr.22): normal alleles: 10–32 repeats; full mutation alleles (expansion): ≥800 ATTCT repeats. Reference sequence: NM_013236.3.
- (CAG)n in intron 2 of the *PPP2R2B* gene (chr.5): normal alleles: 4–32 repeats; full mutation alleles (expansion): >40 CAG repeats. Reference sequence: NM_181678.2.
- (CAG/CAA)n in exon 3 of the *TBP* gene (chr.6): normal alleles: 25–40 repeats; reduced penetrance alleles: 41–48 repeats; full mutation alleles (expansion): >49 repeats. Reference sequence: NM_003194.4.
- (CAG)n in exon 6 of the *ATN1* gene (chr.12): normal alleles: 6–35 repeats; premutation alleles: 36–47 repeats; full mutation alleles (expansion): ≥48 repeats. Reference sequence: NM_001940.3.
- (GAA)n in the first intron of the *FXN* gene (chr.9): normal alleles: 5–33 repeats; premutation alleles: 34–65 repeats; pathological alleles with expansion (full penetrance): >66 repeats. Reference sequence: NM_000144.4.

Sequencing performed by Gencell Genética Avanzada in Bogotá, Colombia in 2021.

4. **Clinical Whole Genome Sequencing**

Sequenced reads were aligned using BWA-mem to GRCh37. Variants were classified and reported using the Golden Helix VarSeq analysis workflow, which implements the guidelines of the *American College of Medical Genetics* (ACMG). The following databases and *in silico* algorithms are used to evaluate and report the impact of variants in the context of human disease: 1000 Genomes, gnomAD, ClinVar, OMIM, dbSNP, NCBI RefSeq Genes, ExAC Gene Restraints, VS-SIFT, VS-PolyPhen2, PhyloP, GERP++, GeneSplicer, MaxEntScan, NNSplice, and PWM Splice Predictor. The analysis was reported using the nomenclature of the *Human Genome Variation Society* (HGVS) (www.hgvs.org/mutnomen) as implemented by the VarSeq transcript annotation algorithm.

Sequencing performed by Gencell Genética Avanzada in Bogotá, Colombia in 2020.

5. **Sanger Sequencing:**

Primers for exon 15 of *PLA2G6* were obtained from Thermo Fisher, while primers for the intronic variants were designed using the online software Primer3 (https://primer3.ut.ee/), considering parameters such as primer length, GC content and product size, to optimize amplification and sequencing quality.

Primer pairs were subsequently standardized by gradient PCR, obtaining optimal annealing temperatures.

a. **P*LA2G6* c.2132C>G (p.Pro711Arg)**

Forward primer: ACACCAAAGGCCCCACAGGAT

Reverse Primer: GTGGTCCTGTTGGTACACAGATG

Chr22 38113398-38113898 501bp amplicon.

Annealing temperature: 65°C

ACACCAAAGGCCCCACAGGATgaggggaagccatcgacctgggctacagaccctgagggaagtggcctgggggag
gggcccacactcacacagtccaccaccatcttgcccagttccttggccccaaaaacagtcttggccagctcccaggggttgctg[g/c]
gacggaagacatccacacaggtcacaggcacttgtggggacctccctgtccccagggagacaacgatggagagtttcttcaccttgtt
ggcctgaccctgttgggaacaggacaggggcagtcagaagagaccttccacaggtagggggcacgaaggggagcgtcaaggg
gaggcccaagactgggctctggggcacatgagaacagggcaagaggtctctggtggcaagagcatgcctggcatgatttccagcc
agcctcttgcacgtgaccccagaacggctggtgcgggcatcacatgccatctgctcaggacagCATCTGTGTACCAACAG
GACCAC

b. **P*LA2G6* c.2034+355G>A**

Forward primer: CAAATTGATCTCACTGGAACCTGT

Reverse Primer: GATCACTGATTCTGCACTGCTC

Chr22 38115044-38115390 347bp amplicon.

Annealing temperature: 57°C

CAAATTGATCTCACTGGAACCTGTtccatgggtgggatctgcctgaagcactgtgctaagggggatctgaggaccact
gtgctctaagtgtgcatgagtggggagggcggcatgcataccatgaacgtgccactcgagccagcctggggacaggacacatgctt
gcccttgagctcacctctgtggatagacagtcccacaggaatgcctgtccccac[g/a]ggttgccggtgctgtggtggaggaccgtga
ggcctctgctaaaaggcctgggggctcggaaagggctccatggaacaggggacccatcctgaggcccttggggacaGAGCAG

TGCAGAATCAGTGATC

Sanger sequencing was performed by GENEWIZ from Azenta and analyzed using the Ugene software v53.1.
